# Community-based Health Insurance Membership Renewal and Associated Factors among Communities in Addis Ababa, Ethiopia

**DOI:** 10.1101/2022.09.06.22278969

**Authors:** Walelign Mekonnen Gashaw, Kidanemariam G/Michael Beyene, Mesafint Abeje Tiruneh

**Affiliations:** Akaki Kality Sub-city Health Office, Addis Ababa, Ethiopia; Ethiopian Food and Drug Authority, Addis Ababa, Ethiopia; Ministry of Heath-Ethiopia, Addis Ababa, Ethiopia

**Keywords:** Community-based health insurance, membership renewal, community, Addis Ababa

## Abstract

**Introduction:** Utilization of contemporary health services in developing nations has remained low. Ethiopia has introduced community-based health insurance (CBHI) to improve access to health services and reduce out-of-pocket payments. However, high dropout and low enrolment rates hamper implementation of CBHI scheme. This study aimed to assess CBHI membership renewal and associated factors among communities in Addis Ababa.

**Methods:** Community-based cross-sectional mixed study design was employed among 626 CBHI members and ten key informants in Addis Ababa from July 05 to August 30, 2021. Multi-stage sampling technique was used to select study participants, and purposive sampling technique was used to select key informants. Quantitative data was collected using structured interviewer administered questionnaire and analyzed using SPSS version 23.0, whereas semi-structured questionnaire was used for in-depth interviews. Binary logistic regression and thematic analysis were employed for quantitative and qualitative data respectively.

**Results:** Membership renewal of the community was 67.3%. Family size (AOR=2.54; 95%CI: 1.26-5.12), knowledge (AOR=1.87; 95%CI: 1.05-3.32), years of enrollment (AOR=2.22; 95%CI: 1.39-3.54), enrollment fee contributor (AOR=9.16; 95%CI: 5.29-15.82), benefit packages (AOR=1.99; 95%CI: 1.18-3.35), trust in CBHI scheme (AOR=3.53; 95%CI: 2.12-5.86), training (AOR=1.81; 95%CI 1.01-3.21), chronic illness (AOR=6.82; 95%CI: 3.80-12.24) and having children under five (AOR=1.98; 95%CI: 1.27-3.10) were significantly associated with CBHI membership renewal. Low level of awareness, limited benefit packages, unaffordable premium, poor quality health services, and shortage of supplies were determinant factors for CBHI membership renewal.

**Conclusion:** CBHI membership renewal was low. Family size, knowledge, years of enrollment, enrollment fee contributor, benefits packages, trust in CBHI scheme, training, chronic illness and having children under five were significantly associated with CBHI membership renewal. In addition, there are correctable gaps in community awareness, benefit packages, affordable premiums, quality health services, and availability of supplies. Hence, necessary measures at all levels must be taken to reduce dropouts from membership.

## Introduction

Universal health coverage (UHC) is becoming a priority. There is a need to increase the financial accessibility of health care services, protect the population from catastrophic expenditure, and decrease the risk of extreme poverty (1). Governments have an increased effort to achieve UHC and to improve healthcare access, use, and financial protection by reducing direct out-of-pocket payments (OOP) for health care services (2, 3). However, many low and middle-income countries are unable to meet their citizens’ healthcare needs. These countries face challenges in raising sufficient funds. Due to limited economic resources and weak government institutions, it is difficult to achieve healthcare financing (4).

Global statistics show that the OOP of health care in low- and middle-income countries is very high. People in these countries are also affected by man-made and natural factors, which further increase medical expenses. This fact is true at the individual, family, and national levels (5).

Because of the unaffordable financial catastrophes, funding the healthcare system is a global concern for all nations (6).

Different forms of community-based health insurance applying the principle of risk-sharing were organized to provide financial risk protection, especially for people in the poor category, to ensure that no one is left behind concerning access to health care services (7). The World Health Organization’s (WHO) view on the CBHI program is that there is a fragile integration between public policy and organizational planning and action (8). Studies indicated a high level of drop-out from the CBHI system, but it has not been analyzed in-depth, and this situation may be a sustainability problem for the CBHI scheme in many low and middle income countries (9, 10).

The CBHI program offers financial protection by reducing out-of-pocket medical expenses and enhancing cost recovery. However, low enrolment rates hamper the successful development of CBHI schemes and endanger the sustainability of CBHI schemes not only because they reduce the size of the insurance pool but also because they bear a negative impact on subsequent enrolment and dropout (11).

In Ethiopia, CBHI schemes have been introduced to reduce the financial shock due to unexpected and irregular out-of-pocket payments and to increase the resource pooling for healthcare that would improve access and utilizations of health services (12, 13) The government of Ethiopia depends heavily on outdoor supporters and out-of-pocket payments to fund health services for its citizens (13, 14)

The government of Ethiopia introduced the CBHI scheme to address the problems of OOP expenses and increase enrollment of the community residing in rural and urban. This is an ingredient of the health care financing reform strategy that aims to recover quality and access to health services, and remove financial burdens (15).

CBHI implementation began in 2010/11 as pilot schemes in 13 districts of Amhara, Oromia, Southern Nations Nationalities and Peoples (SNNP), and Tigray regional states and expanded CBHI schemes to 161 districts after three years of piloting (15, 16).

In addition, Addis Ababa in all its sub cities is one of the CBHI implementing regions starting January 2018 (17). According to the Ethiopian health insurance report, the 2019 enrollment rate in Addis Ababa was 80% which is higher as compared to the national report (44%). The 2020/21 nine-month reports of Ethiopian health insurance in Addis Ababa showed that the enrollment rate decreased by 5% (18). Identifying determinant factors of CBHI membership renewal would help in devising strategies that promote communities’ membership renewal. Hence, the study aimed at assessing membership renewal and determinant factors of renewal of CBHI membership in Addis Ababa.

## Materials and Methods

### Study design, setting and period

A community-based cross-sectional mixed study design method was employed with sequential explanatory design to triangulate the quantitative data with the qualitative data. The data was collected in Addis Ababa from July 05 to August 30, 2021.

Addis Ababa is the diplomatic capital of the African Union and the capital city of Ethiopia. It has 11 sub-cities and 117 districts. The city has an estimated population of 3.7 million, of which 53.6% are females and 46.4% are males (19, 20). Based on data obtained from the Ministry of Health and Addis Ababa Health Bureau, there were 22 public hospitals, 102 health centers, and 1327 clinics in Addis Ababa. Furthermore, there were an estimated 960,676 households in Addis Ababa. Among these, 231,677 households were enrolled in the CBHI scheme (17).

### Source and study population

The source population was all CBHI scheme members living in Addis Ababa, and selected CBHI member households who resided in the selected sub-cities in Addis Ababa were the study population. The key informants for the in-depth interviews were CBHI core process coordinators, medical directors, and CBHI members living in Addis Ababa. Members of CBHI whose enrollment status was less than a year and households that used a fee waiver scheme were excluded from the study.

### Sample size determination

The sample size was determined by using the single population proportion formula for the CBHI membership renewal and the double population proportion formula for predictor variables, assuming a 25% proportion of renewed CBHI membership taken from a previous study done in Gimbichu district, Oromia Region (21) at a 95% confidence interval, margin of error of 5% (0.05), design effect of 2 and a 10% non-response rate. Accordingly, a total sample size of 634 households was taken. The sample size for the qualitative study was guided by the degree of information saturation based on preliminary analysis during data collection. Hence, ten key informants from different positions and members were selected for the in-depth interviews.

### Sampling procedure

The study participants were drawn using a multi-stage sampling technique. Three sampling stages were used to select the final study participants. In the first stage, four sub-cities were selected randomly using a lottery method out of eleven sub-cities (40% of the sub-cities). The second stage involved the selection of districts from four sub-cities. For each selected sub-city, the number of districts was listed, and a simple random sampling technique using a lottery method was used to select districts in each selected sub-city considering the number of districts. Finally, a list of households was obtained from each district’s administration offices, which was used as a sampling frame. The sample size was proportionally allocated to selected districts based on each district’s number of households. Then, a simple random sampling technique (lottery method) was employed to select the households (30% of the households from the selected districts).

A purposive sampling technique was used to select the key informants for the qualitative study. The key informants were selected from CBHI scheme, hospitals, health centers, and community members who have better knowledge of the program.

### Data collection procedures and quality assurance

An interviewer-administered structured questionnaire adapted from previous literature and tailored to the study was used to collect quantitative data. The questionnaire contains socio-demographic characteristics, knowledge factors, scheme-related factors, and health service-related variables. The questionnaire was originally prepared in English, translated into Amharic, and then back to English to check for its consistency of meaning.

A one day training was given to data collectors and supervisors. The training was emphasized on the purpose of the study; data collection tools; data collection procedure; principles and ethical considerations; and the importance of privacy and confidentiality of respondents. Regular supervision was given during data collection. The collected data was evaluated by supervisors and the primary investigator before data entry on a daily basis for data clarity, completeness, consistency, and uniformity. The data collection tool was pretested on 5% of randomly selected households from non-selected districts, which were not included in the study. Accordingly, the questionnaire was slightly modified.

A one-on-one, face-to-face, in-depth interview was carried out with key informants using a semi-structured, open-ended interview questionnaire with flexible probing techniques to elaborate on the original responses of the interviewee. All interviews were conducted in the key informant’s preferred language (Amharic or English), and audio was recorded with the permission of the interviewee and notes were taken properly by the interviewer. All interviews were held in private locations to express an idea well. The audio recording was transcribed verbatim and translated into English by local native speakers. To check the accuracy of the translation, one of the recordings was translated and transcribed by a bi-lingual expert and compared with the primary work. Furthermore, the findings of the study were communicated to the supervisor of the key informants for the authenticity of the transcripts and interpretations. In addition, pilot interviews were carried out to check any practical problems that may encounter the researcher during actual interview periods and modify them accordingly.

### Data management and analysis

The collected quantitative data was entered into Epi-Info version 7.2.1.0 and then exported to SPSS version 23.0 for analysis. The collected data was cleaned for anomalies by running frequencies and cross-tabulations after exporting to SPSS. Erroneous data was cross-checked with the hard copies of the completed questionnaires. Descriptive statistics were computed for each variable and presented using frequencies, percentage, narrations and tables. At 25% level of significance, univariate binary logistic regression analysis was done for each variable to identify the potentially significant independent variables. Variables that had a p-value < 0.25 on univariate logistic regression analysis were entered into multivariable logistic regression analysis.

The association between the dependent variable and independent variables was analyzed using a binary logistic regression analysis model. A p-value<0.05 was considered as statistically significant. The strength of association and precision were examined using an adjusted odds ratio at a 95% confidence interval. To check the adequacy of the final model Hosmer-Lemeshow goodness of fit test was checked and the model fitted to the data and multi-collinearity was checked and no multi-collinearity detected.

The audio records and notes of the interviews were transcribed using a verbatim transcription technique. The transcribed scripts were intensively read, and the data were categorized into themes. Thematic analysis method was used to analyze the qualitative data. The analysis was facilitated using Open Code version 4.0.2.3 software. The results of the qualitative study were presented in narration, quotations and triangulated with quantitative results. In reporting findings, codes were used to maintain anonymity of the key informants.

### Ethical consideration

Ethical approval was sought from GAMBY Medical and Business College (GAMBY, IRERC, 2021) and Addis Ababa Health Bureau Research Ethics Review Committees (Ref. No: A/A/H/323/227). In addition, a permission letter was sought from Addis Ababa Regional Health Bureau, followed by sub-city health offices where the study site is located. The purpose of the study was described to study participants. All study participants were also informed about their right of not participating in the study at any time and written informed consent was obtained from the study participants. The confidentiality of information and the privacy of participants during the interview were respected. A detailed explanation was given to the study participants about the objectives and benefits of the study.

## Results

### Socio-demographic characteristics of the study participants

Out of the 634 members of CBHI, 626 of them participated in the study making the response rate of 98.7%. The majority of the study participants, 365 (58.3%), were females. The mean age of the respondents was 41.5 (SD: ±11.17) years, ranging from 20–80 years, and 351 (56.1%) of them were in the age group of 20–40 years. Five hundred and sixty (89.5%) of the study participants were married. About 169 (27%) of the study participants had completed primary (1–8th) school, and 167 (26.7%) of them were housewives. Five hundred and thirty households (84.7%) had a family size of five or fewer family members, and 289 (46.2%) of the study participants had a monthly average family income of 1600–4000 ETB **(Table 1)**.

**Table 1:** Socio-demographic characteristics of the study participants in Addis Ababa, Ethiopia 2021 (n=626)

| Variables | Frequency (n) | Percent (%) |
| --- | --- | --- |
| **Gender** |  |  |
| Male | 261 | 41.7 |
| Female | 365 | 58.3 |
| **Age in years** |  |  |
| 20-40 | 351 | 56.1 |
| 41-59 | 207 | 33.1 |
| $\geq 60$ | 68 | 10.9 |
| **Marital status** |  |  |
| Not married | 66 | 10.5 |
| Married | 560 | 89.5 |
| **Educational status** |  |  |
| Illiterate | 70 | 11.2 |
| Read and write only | 116 | 18.5 |
| Primary | 169 | 27.0 |
| Secondary | 147 | 23.5 |
| Certificate and diploma | 93 | 14.9 |
| Degree and above | 31 | 5.0 |
| **Occupation** |  |  |
| Daily labor | 101 | 16.1 |
| Housewife | 167 | 26.7 |
| Merchant | 97 | 15.5 |
| Government employee | 14 | 2.2 |
| Private organization | 138 | 22.0 |
| No job | 109 | 17.4 |
| **Family size** |  |  |
| ≤5 | 530 | 84.7 |
| >5 | 96 | 15.3 |
| **Monthly income** |  |  |
| <1600 | 200 | 31.9 |
| 1600-4000 | 289 | 46.2 |
| >4000 | 137 | 21.9 |

### Knowledge, experiences and expectations of CBHI Members

Among the total study participants, 517 (82.5%) of households had adequate knowledge about the CBHI scheme. About 221 (35.3%) of study participants were enrolled for about two or more years. More than one-third, 236 (37.7%) of the CBHI members got information about CBHI services from community organizers. Out of 626 study participants, 140 (22.4%) of them were paid by the support of others.

The contribution fee is affordable for the community, according to the majority of study participants, 349 (55.8%). About 424 (67.7%) of the study participants also agreed that the CBHI benefit packages satisfy the needs of households. Almost half of study participants (318/626, 50.8%) believed that contracted health facilities provide quality healthcare services to insured households. Four hundred-seventeen (66.6%) of CBHI members trust the CBHI scheme that it provides medical services they need with the contribution they make, and 367 (58.6%) of the study participants agreed that district CBHI officials provide good service during ID preparation and premium collection. Only 117 (18.7%) of the study participants were participating in CBHI related training **(Table 2)**.

**Table 2:** Experience and expectations of the study participants in Addis Ababa, Ethiopia, 2021 (n = 626).

| Variables | Frequency (n) | Percent (%) |
| --- | --- | --- |
| **Enrolment year** |  |  |
| <2 | 405 | 64.7 |
| ≥2 | 221 | 35.3 |
| **Information source** |  |  |
| District health professionals | 182 | 29.1 |
| Community organizers | 236 | 37.7 |
| Health professionals in the health facility | 80 | 12.8 |
| Neighbors/friends | 74 | 11.8 |
| Mass media such as TV/Radio | 54 | 8.6 |
| **Contribution fee contributor** |  |  |
| My self | 486 | 77.6 |
| With help | 140 | 22.4 |
| **Affordability of premium** |  |  |
| Strongly agree | 94 | 15 |
| Agree | 349 | 55.8 |
| Neutral | 45 | 7.2 |
| Disagree | 127 | 20.3 |
| Strongly disagree | 11 | 1.8 |
| **Benefit packages that satisfy the needs of households** |  |  |
| Strongly agree | 53 | 8.5 |
| Agree | 424 | 67.7 |
| Neutral | 41 | 6.5 |
| Disagree | 95 | 15.2 |
| Strongly disagree | 13 | 2.1 |
| **Quality health service provision** |  |  |
| Strongly agree | 15 | 2.4 |
| Agree | 318 | 50.8 |
| Neutral | 71 | 11.3 |
| Disagree | 211 | 33.7 |
| Strongly disagree | 11 | 1.8 |
| **Trust in CBHI scheme** | 417 | 66.6 |
| Yes | 209 | 33.4 |
| No |  |  |
| **District CBHI official provide good service** |  |  |
| Strongly agree | 182 | 29.1 |
| Agree | 367 | 58.6 |
| Neutral | 15 | 2.4 |
| Disagree | 60 | 9.6 |
| Strongly disagree | 2 | 0.3 |
| **Participating in CBHI training** |  |  |
| Yes | 117 | 18.7 |
| No | 509 | 81.3 |

### Household characteristics and health service provision

About 69 (11%) of the study participants had frequent illness or injury, and 180 (28.8%) of the study participants had a chronic patient in the household. Two hundred and eighty-eight (46%) of the study participants had a child under five years old, and 151 (24.1%) of the households had family members who were over 65 years old. Three hundred and eighty-seven (61.8%) of the total study participants believed that service providers provide equal health care to CBHI members and non-members. Only 257 (41%) of the study participants agreed that medicines were available at the health facilities. More than half, (55.3%) of the study participants agreed on the availability of laboratory services in the health facilities. About 495 (79.1%) of the study participants agreed that health professionals are respectful, and 345 (55.15%) of the study participants were satisfied with the health services provided **(Table 3)**.

**Table 3:**
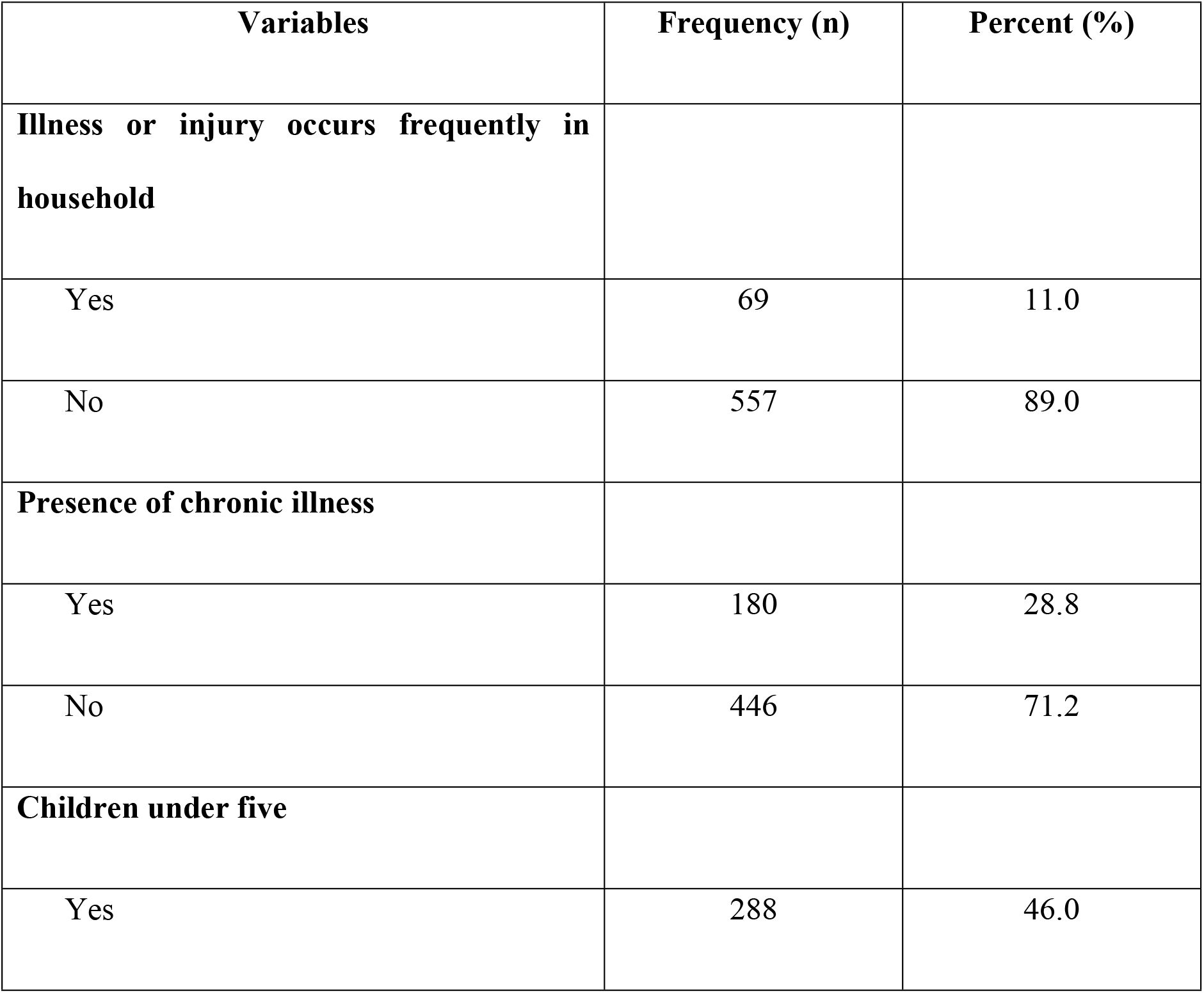
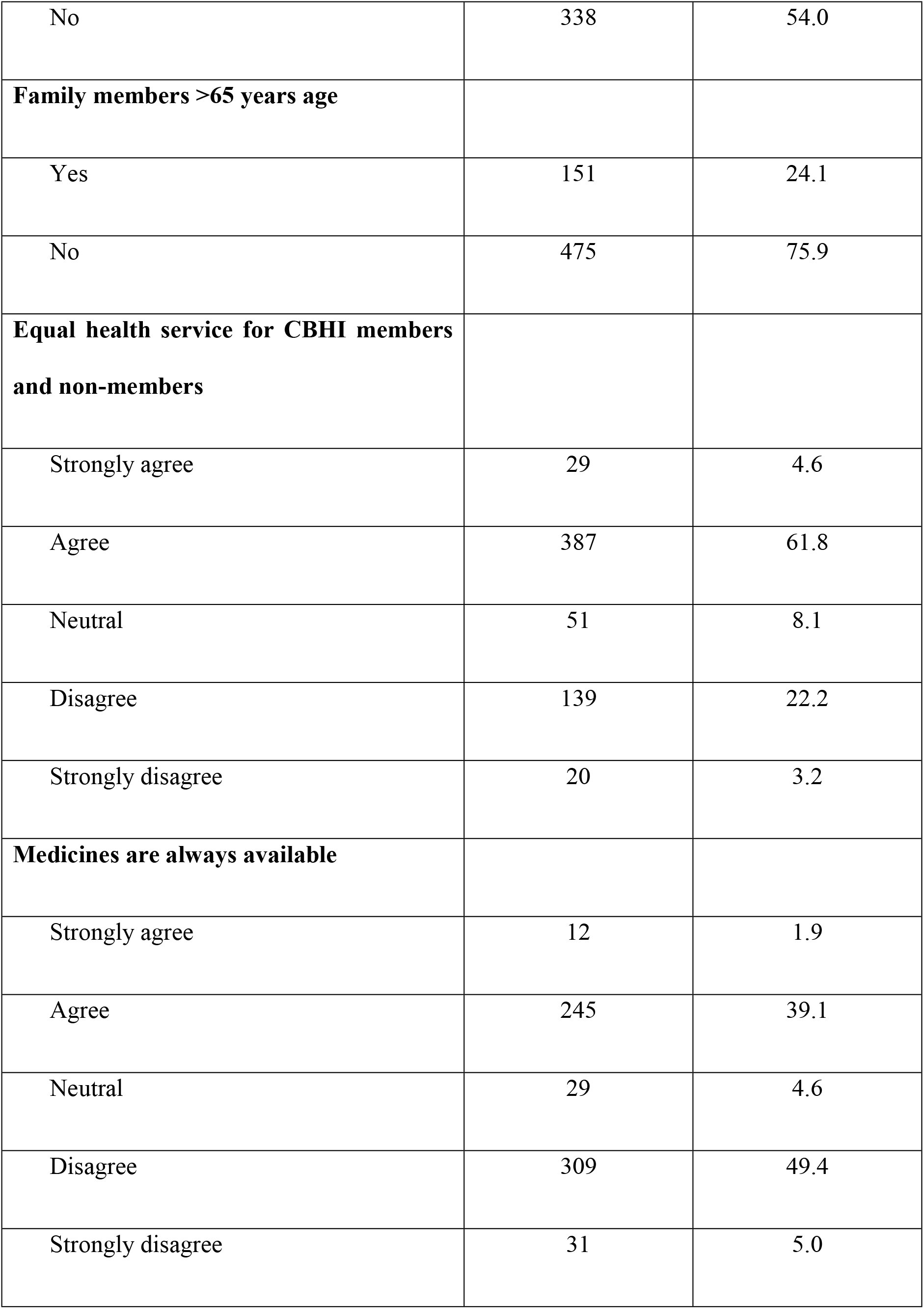
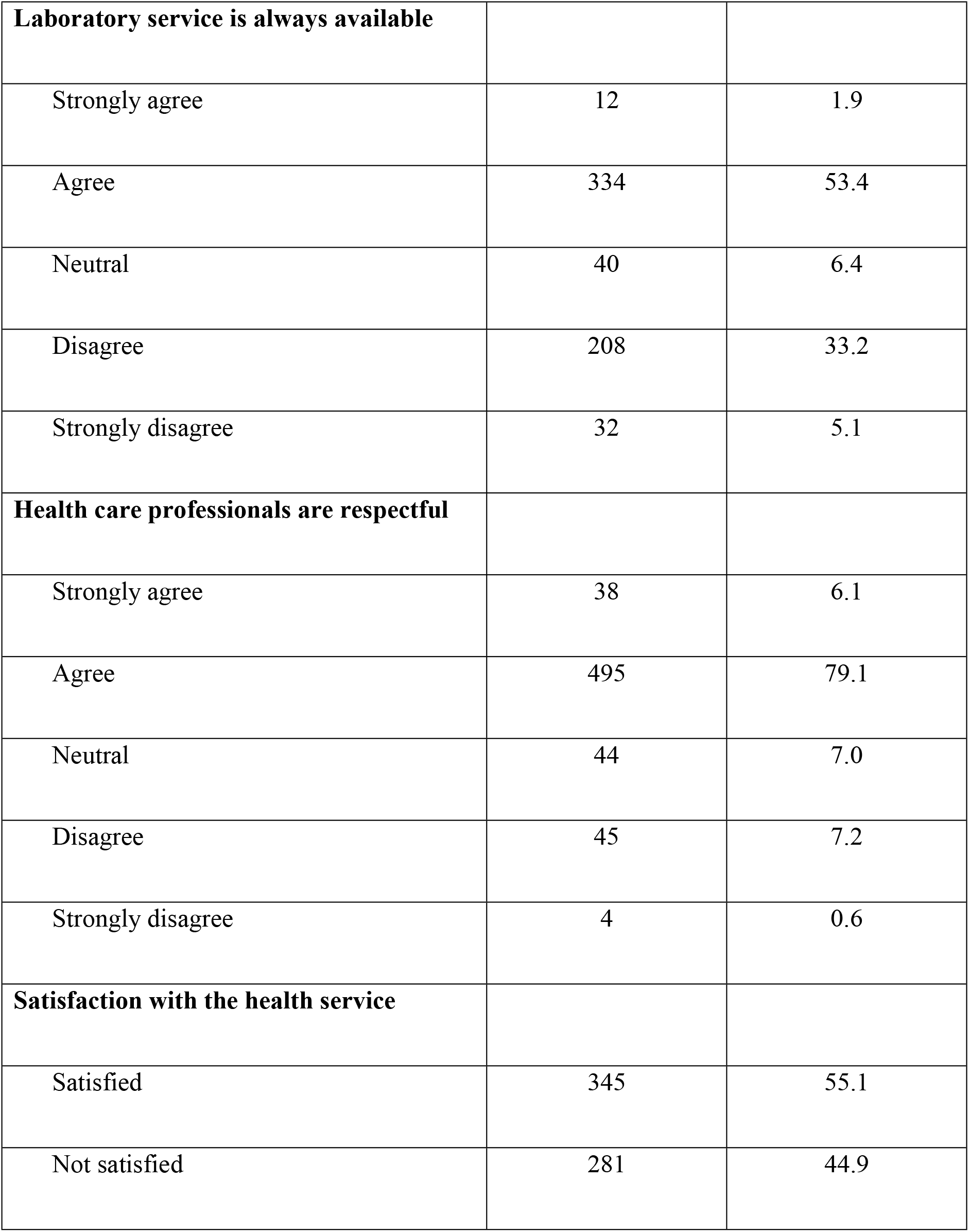
Household characteristics and health service provision among the study participants in Addis Ababa, Ethiopia, 2021 (n = 626).

### Community-based health insurance membership renewal

Among the total study participants, 421 (67.3%) of them renewed their CBHI membership and the remaining 205 (32.7%) of them did not renew their CBHI membership.

### Factors affecting renewal of community-based health insurance membership

In bivariate logistic regression analysis (P<0.25); family size, knowledge about CBHI, years of enrollment, enrollment fee contributor, benefits package, perceived quality of healthcare service, trust in CBHI scheme, training on CBHI, presence of chronic illness, having children under five, availability of medicines, and availability of laboratory services or supplies were found to be significantly associated with CBHI membership renewal.

However, in multivariable binary logistic regression analysis, only family size, knowledge about CBHI, years of enrollment, enrollment fee contributor, benefits packages, trust in CBHI scheme, training on CBHI, presence of chronic illness and having children under five years were significantly associated with CBHI membership renewal.

The odds of CBHI membership renewal among households who had a family size of greater than five was 2.54 times the odds of households who had family size of five or below (AOR=2.54; 95% CI: 1.26-5.12).

The odds of CBHI membership renewal among knowledgeable households were 1.87 times the odds of households who were not knowledgeable about CBHI (AOR=1.87; 95% CI: 1.05-3.32).

The odds of CBHI membership renewal among households who were enrolled for two or more years was 2.22 times the odds of households who enrolled for less than two years (AOR=2.22; 95% CI: 1.39-3.54).

The odds of CBHI membership renewal among households who covered their fee by themselves was 9.16 times the odds of households who covered their fee by the help of others (AOR=9.16; 95% CI: 5.29-15.82).

The odds of CBHI membership renewal among households who agreed that the benefit packages fulfill the needs of the households was 9.16 times the odds of those households who disagreed (AOR=1.99; 95% CI: 1.18-3.35).

The odds of CBHI membership renewal among households who had trust on CBHI scheme were 3.53 times the odds of households who had no trust on CBHI scheme (AOR=3.53; 95% CI: 2.12-5.86).

The odds of CBHI membership renewal among households who participated in CBHI training was 1.81 times the odds of households who did not participate (AOR=1.81; 95% CI 1.01-3.21).

The odds of CBHI membership renewal among households who had a family member with chronic illness was 6.82 times the odds of households who had no family member with chronic illness (AOR=6.82; 95% CI: 3.80-12.24). Moreover, the odds of CBHI membership renewal among households who had under-five children was 1.98 times the odds of households who had no under-five children (AOR=1.98; 95% CI: 1.27-3.10) **(Table 4)**.

**Table 4:** Factors associated with community-based health insurance membership renewal among the community in Addis Ababa, 2021 (n=626).

| Variable | Membership renewal |  | COR (95% CI) | AOR (95% CI) | p-value |
| --- | --- | --- | --- | --- | --- |
|  | Yes | No |  |  |  |
| Family size |  |  |  |  |  |
| ≤5 | 341 | 189 | 1.00 | 1.00 |  |
| >5 | 80 | 16 | 2.77 (1.57-4.88) | 2.54 (1.26-5.12) | 0.009* |
| Knowledge about CBHI |  |  |  |  |  |
| Knowledgeable | 378 | 155 | 2.84 (1.81-4.44) | 1.87 (1.05-3.32) | 0.033* |
| Not knowledge | 43 | 50 | 1.00 | 1.00 |  |
| Years of enrollment |  |  |  |  |  |
| <2 | 254 | 151 | 1.00 | 1.00 |  |
| ≥2 | 167 | 54 | 1.84 (1.27-2.65) | 2.22 (1.39-3.54) | 0.001* |
| Enrollment fee contributor |  |  |  |  |  |
| Household | 367 | 119 | 4.91 (3.30-7.31) | 9.16 (5.29-15.82) | <0.001* |
| Others (help) | 54 | 86 | 1.00 | 1.00 |  |
| Benefit packages fulfill needs of households |  |  |  |  |  |
| Agreed | 345 | 132 | 2.51 (1.72-3.67) | 1.99 (1.18-3.35) | 0.010* |
| Disagreed | 76 | 73 | 1.00 | 1.00 |  |
| Households get access to quality healthcare services |  |  |  |  |  |
| Agreed | 259 | 74 | 2.83 (2.00-4.00) | 1.37 (0.83-2.27) | 0.221 |
| Disagreed | 162 | 131 | 1.00 | 1.00 |  |
| Trust in CBHI scheme |  |  |  |  |  |
| Yes | 323 | 94 | 3.90 (2.73-5.56) | 3.53 (2.12-5.86) | <0.001* |
| No | 98 | 111 | 1.00 | 1.00 |  |
| Household participated in<br>CBHI training |  |  |  |  |  |
| Yes | 90 | 27 | 1.79 (1.12-2.86) | 1.81 (1.01-3.21) | 0.045* |
| No | 331 | 178 | 1.00 | 1.00 |  |
| Household member<br>has chronic illness |  |  |  |  |  |
| Yes | 150 | 30 | 3.23 (2.10-4.99) | 6.82 (3.80-12.24) | <0.001* |
| No | 271 | 175 | 1.00 | 1.00 |  |
| Household with children<br>under five |  |  |  |  |  |
| Yes | 211 | 77 | 1.67 (1.19-2.35) | 1.98 (1.27-3.10) | 0.003* |
| No | 210 | 128 | 1.00 | 1.00 |  |
| Medicines always available in the health facility |  |  |  |  |  |
| Agreed | 185 | 72 | 1.45 (1.03-2.05) | 1.35 (0.83-2.20) | 0.224 |
| Disagreed | 236 | 133 | 1.00 |  |  |
| Laboratory services always available in health facility |  |  |  |  |  |
| Agreed | 252 | 94 | 1.76 (1.26-2.47) | 1.50 (0.96-2.35) | 0.078 |
| Disagreed | 169 | 111 | 1.00 | 1.00 |  |

*Note: COR: crude odds ratio; AOR: adjusted odds ratio; CI: confidence interval; \*statistically significant at 5% level of significance*

### Qualitative findings

A total of ten in-depth interviews were conducted with the key informants. The key informants were three medical directors, three CBHI team leaders, and four CBHI members. Their ages ranged from 30 to 57 years.

The key informants were asked about the overall renewal, benefit packages, availability of services and supplies, and related issues about CBHI membership renewal. Key themes emerged from the analysis were (i) Member’s awareness about CBHI; (ii) affordability; (iii) health care service delivery; (iv) Availability of supplies; and (v) reasons for dropout. Analysis of the qualitative data from interviews with key informants is presented in detail as follows.

### CBHI member’s awareness

Most of the key informants explained that awareness training on health insurance is not being provided in either district or sub-city. The members of the CBHI scheme often did not clearly understand the services to be provided and not to be provided by the scheme. One of the key informants mentioned that:

*“…*.. *the CBHI scheme office did not provide adequate awareness on how, when, and where to use services. Members claim without understanding their rights and responsibility. CBHI officials more concerned with increasing members without creating awareness” (HI01)*.

### Affordability of premium

Many key informants said that membership contributions were affordable and do not affect the community, but some oppose this. One of the key informants raised about the need for an income-based payment system. This was underlined as shown below:

*“…. It is difficult to say that the membership fee is reasonable because there are several groups of the society that cannot pay 370 ETB and those with better incomes can pay more. Therefore, there is no income-based payment system. it is good to make the contribution based on the income of the households” (HI02)*.

Another participant who dropped from the scheme also mentioned that:

*“…*.. *I became a member of the CBHI scheme last year. Even though the contribution fee was not difficult with the help of someone else, I did not renew my health insurance because I could no longer afford it” (HI03)*.

### Service delivery for CBHI members

Most of the service providers and members of the CBHI believed that there are several problems with service delivery by health facilities and professionals. This was supported by one of the participant that:

*“…*.. *In the health facility, CBHI members are not treated equally as non-members were especially in the pharmacy room if members of CBHI and non-member of CBHI come to buy aq drug at the same time; nonmembers will get the service first because they pay directly from out of pocket and the CBHI members consider that health professionals have a negative attitude for members because we are not paid directly from out of pocket” (HI08)*.

### Availability of supplies

The key informants thought that lack of supply of medicines, supplies and other resources made them dissatisfied and not continued with the scheme. The problem is not resolved at the level of health facilities. This was supported by one of the participants:

*“…*.. *We do not have an adequate supply of medicines and laboratory reagents and supplies because of the interruption of the supply of materials from the government supplier and there is a lack of budget to buy from private suppliers” (HI07)*.

Another key informant supported the unavailability of the medicines and supplies. But, one of the participants mentioned that there is another option for CBHI members to access medicines in Addis Ababa. She pointed out that:

*“…*.. *Adequate medicines and laboratory supplies are not available in public health facilities. However, Kenema pharmacies in Addis Ababa offered medicines without branding. But there is no alternative to laboratory services except for the private ones” (HI10)*.

### Reasons for dropout of members

The key informants argued that despite the service delivery problems, many members of the community are using it. However, members are terminating their membership for various reasons. This was supported by one of the participant that:

*“…*.. *The main reasons for members to drop out were the poor quality of health services, limited benefit packages compared to needs of members, members not knowing the renewal period and the severe shortage of pharmaceuticals, laboratory supplies and radiological devices. In addition, I do not think the benefit framework fully meets the needs of the members, especially brand drugs” (HI 09)*.

Furthermore, another participant mentioned that the current health services provision must be improved before all: He augmented this concern as follows:

*“*………. *the current quality of health services is not optimal; there are challenges in receiving timely healthcare services. Again, we contributed a fee to get appropriate services but from my experience medicines are frequently out of stock in public facilities. That means we will be forced to purchase from private institutions that are not part of the scheme. It is frustrating while you are contributing but getting nothing. It is better to improve the quality and availability of services before implementing the scheme. Otherwise it might fail and it ultimately erodes public trust and will have unexpected consequences” (HI06)*.

## Discussion

The study assessed CBHI membership renewal and associated factors among the community in Addis Ababa. The study found that the CBHI membership renewal rate of the community was 67.3%. The CBHI membership renewal rate of the community in our study was lower when compared with the national renewal rate (82%) in April 2013 (11) and much greater than a study conducted in Gimbichu district, Oromia Region (21) and India (22). The possible reasons for the difference might be due to differences in study methods, health care settings, and awareness of the community to CBHI scheme.

The odds of CBHI membership renewal among households who had a family size of greater than five was 2.54 times the odds of households who had family size of five or below. This finding was supported by one of the systematic reviews in lower and middle-income countries (23) and the study conducted in the West Gojjam Zone (24). A positive association between family size and CBHI enrollment and renewal could be a result of the financial burden that large households face when attempting to pay for health care out of pocket.

The odd of CBHI membership renewal among knowledgeable households was 1.87 times the odds of households who were not knowledgeable about CBHI. This finding is supported by the qualitative study. The qualitative finding indicated that its members often do not clearly understand the services that are not provided by the scheme. Due to this reason households might not renew their membership.

The odds CBHI membership renewal among households who was enrolled for two or more years was 2.22 times the odds of households who enrolled for less than two years. The finding of this study is comparable with studies conducted in India (22) and Yirgalem, Southern Ethiopia (25). The possible justification might be households who enrolled for a longer period of time well understand the advantage of community based health insurance and need to keep their membership than those who enrolled for a shorter period of time and willing to keep into the scheme (26).

The odds of CBHI membership renewal among households who covered their fee by themselves was 9.16 times the odds of households who covered their fee by the help of others. This finding is supported by the qualitative findings. Households mentioned affordability as one of the determinant factors to renew membership and they did not renew their membership because they were unable to cover the required fee.

The odds of CBHI membership renewal among households who agreed that the benefit packages fulfill the needs of the households was 9.16 times the odds of those households who disagreed. This finding is in line with the qualitative finding. Limited benefit package was identified as one of the determinant factors not to renew membership. The households believed the benefit package of the scheme is limited and it does not fulfill their need.

The odds of CBHI membership renewal among households who had trust in the CBHI scheme were 3.53 times the odds of households who had no trust in the CBHI scheme. Similarly, the thematic synthesis in low and middle-income countries found that trust in the scheme and its management was a significant enabler of renewal (23).

The odds of CBHI membership renewal among households who participated in CBHI training was 1.81 times the odds of households who did not participate. It was also supported by the qualitative findings of this study. Lack of awareness on the benefit of health insurance leads to risk of dropout from the scheme. This was complemented with another study conducted in Tigray Region, the case of Kilte Awlaelo district (27).

The odds of CBHI membership renewal among households who had a family member with chronic illness was 6.82 times the odds of households who had no family member with chronic illness. This study was in line with the study done in West Shewa Zone, Oromia Region (28). Since poor health status typically requires greater health care utilization, it is possible that households with sicker members were more likely to renew or enroll because they could obtain desired health services through the CBHI scheme.

The odds of CBHI membership renewal among households who had under-five children was 1.98 times the odds of households who had no under-five children. This finding is supported by the study done in Burkina-Faso (29). The possible justification might be that under-five children need more health care and they are a concern of a household and it might be difficult to cover the health care expense by out of pocket expenditure.

Many other factors were identified which affect CBHI membership renewal by the qualitative study. Affordability premium and insurance benefit packages were among the most frequently raised issues. But few participants strongly opposed the inclusion of expensive services like cosmetic surgery, dialysis and eyeglass provision fearing that covering such services would drain the fund thereby compromising universal health coverage.

In addition, quality of health services in health facilities was an important issue found during interviews and poor quality health service mentioned as one of the determinant factors for membership renewal. Majority of the participants were not satisfied with the availability of health services, medicines and supplies.

### Strengths and limitations

The strength of the study was that the quantitative data was triangulated with qualitative findings. Furthermore, the study considered factors such as knowledge, trust of the CBHI scheme, quality of health service, district health officials’ service provision, equality for members and non-members in health service provision, and affordability. The study had some limitations as well. The study findings might be affected by social desirability bias among the community. In addition, the study might have had response bias among key informants.

### Conclusions

CBHI membership renewal was low. Family size, knowledge, years of enrollment, enrollment fee contributor, benefits package, trust on CBHI scheme, training, presence of chronic illness and having children under five were significantly associated with CBHI membership renewal. In addition, low level of awareness, limited benefit packages, unaffordability of premium, perception of poor quality of service provision, and shortage of supplies were cited by key informants as factors affecting CBHI membership renewal. Hence, necessary measures should be taken at all levels to increase membership renewal.

## Data Availability

All data produced in the present work are contained in the manuscript

## Declarations

### Consent to publish

The consent for publication was obtained from each study participants during data collection.

### Availability of data and materials

The data set supporting the conclusions of this article are available in the manuscript.

### Authors’ contributions

Both authors meet the ICMJE criteria for co-authorship, providing substantial intellectual contributions for the manuscript. WMG conceived the idea, wrote the proposal, participated in the data collection process, analyze data and draft the manuscript. KGB approved the proposal with some revisions, participated in data analysis and interpretation of data, and reviewed the manuscript. MAT participated in data analysis and interpretation of data, reviewed and finalized the manuscript. All authors approved the final draft of the manuscript and agree to be accountable for all aspects of the work in ensuring that questions related to the accuracy or integrity of any part of the work are appropriately investigated and resolved.

## Acknowledgments

The authors would like to thank GAMBY Medical and Business College, Addis Ababa City Health Bureau, Sub-Cities, data collectors, supervisors and study participants for their collaboration and contribution.

